## Supplementary Figures and Tables for "Dynamics of B-cell repertoires and emergence of cross-reactive responses in COVID-19 patients with different disease severity"

Montague *et. al.*

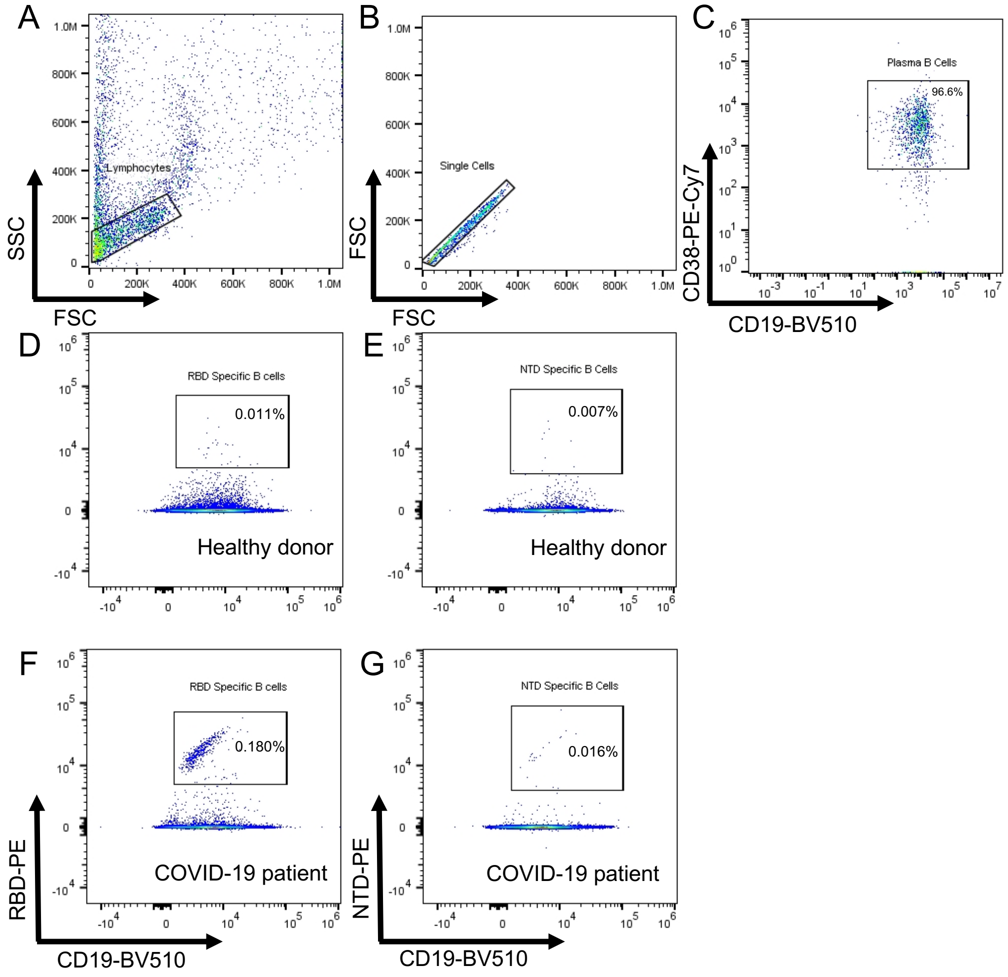


**Figure S1. Gating strategy for the CD38+ Plasma B cell. (A-C)** Flow cytometry stainings of CD38+ plasma B cells from CDVID-19 infected patient using two fluorescent markers, anti-human CD19-BV510 (BioLegend) and CD38-PE-Cy7 (BioLegend) in the same tube. Percentages indicate the proportions of $CD19^{+}CD38^{+}$ plasma B cells within total B cells. **(D-G)** Flow cytometry stainings of RBD- or NTD-specific B cell from healthy donor and CDVID-19 infected patient using anti-human CD19-BV510 (BioLegend) and PE fluorescent dye for RBD protein (D, F) or NTD protein (E, G) in the same staining tube. Percentages indicate the proportions of $CD19^{+}$ and RBD- or NTD-protein double positive specific B cells.

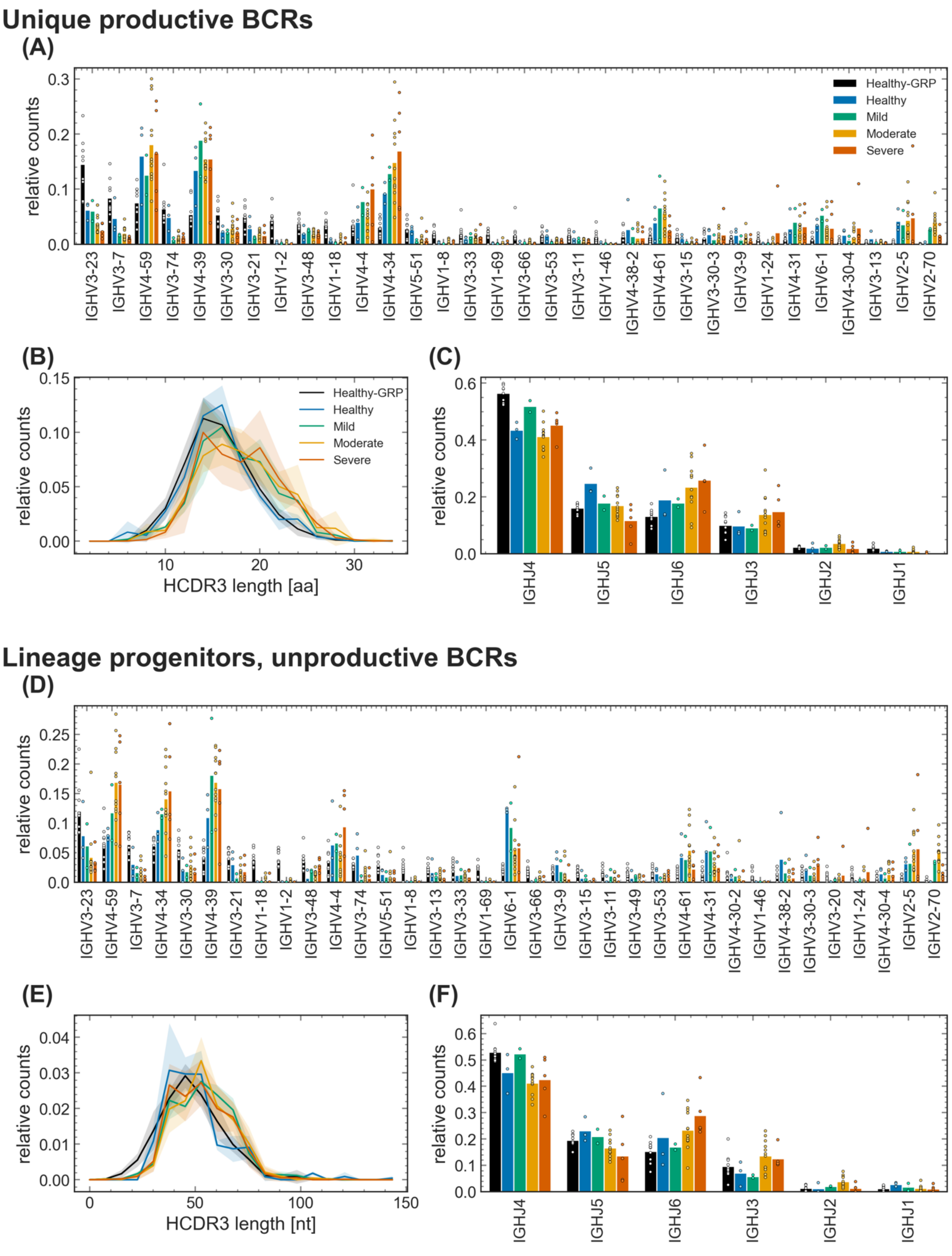

Unique productive BCRs
(A)
relative counts
Healthy-GRP
Healthy
Mild
Moderate
Severe
IGHV3-23 IGHV3-7 IGHV4-59 IGHV3-74 IGHV4-39 IGHV3-30 IGHV3-21 IGHV1-2 IGHV3-48 IGHV1-18 IGHV4-4 IGHV4-34 IGHV5-51 IGHV1-8 IGHV3-33 IGHV1-69 IGHV3-66 IGHV3-53 IGHV3-11 IGHV1-46 IGHV4-38-2 IGHV4-61 IGHV3-15 IGHV3-30-3 IGHV3-9 IGHV1-24 IGHV4-31 IGHV6-1 IGHV4-30-4 IGHV3-13 IGHV2-5 IGHV2-70
(B)
relative counts
HCDR3 length [aa]
(C)
relative counts
IGHJ4 IGHJ5 IGHJ6 IGHJ3 IGHJ2 IGHJ1
Lineage progenitors, unproductive BCRs
(D)
relative counts
IGHV3-23 IGHV4-59 IGHV3-7 IGHV4-34 IGHV3-30 IGHV4-39 IGHV3-21 IGHV1-18 IGHV1-2 IGHV3-48 IGHV4-4 IGHV3-74 IGHV5-51 IGHV1-8 IGHV3-13 IGHV3-33 IGHV1-69 IGHV6-1 IGHV3-66 IGHV3-9 IGHV3-15 IGHV3-11 IGHV3-49 IGHV3-53 IGHV4-61 IGHV4-31 IGHV4-30-2 IGHV1-46 IGHV4-38-2 IGHV3-30-3 IGHV3-20 IGHV1-24 IGHV4-30-4 IGHV2-5 IGHV2-70
(E)
relative counts
HCDR3 length [nt]
(F)
relative counts
IGHJ4 IGHJ5 IGHJ6 IGHJ3 IGHJ2 IGHJ1

**Figure S2. Bulk repertoire sequence statistics. (A-C)** Similar statistics are shown as in Fig. 3 (A, C-D), but for unique receptors excluding singletons (Methods). Unique BCRs in healthy individuals (our control and the Great Repertoire Project (GRP) by (Briney et al., 2019) show significantly shorter HCDR3s compared to moderate and severe cohorts. ANOVA statistics for mean HCDR3 length between cohorts: Healthy-Mild: $F_{1,3} = 8.7$, p-value = 0.06; Healthy-Moderate: $F_{1,13} = 17.2$, p-value = 0.001; Healthy-Severe: $F_{1,6} = 10.0$, p-value = 0.020; GRP-Mild: $F_{1,10} = 11.3$, p-value = 0.0073; Healthy-GRP: $F_{1,11} = 0.074$, p-value = 0.791; GRP-Moderate: $F_{1,20} = 34.0$, p-value = 0.000011; GRP-Severe: $F_{1,13} = 41.5$, p-value = 0.000022. **(D-F)** Similar statistics are shown as in Fig. 2 (A, C-D), but for unproductive lineage progenitors. The differences in the statistics of HCDR3 length between the unproductive repertoires of healthy individuals and the COVID-19 cohorts are insignificant (ANOVA p-value > 0.01). Colors are consistent across panels.

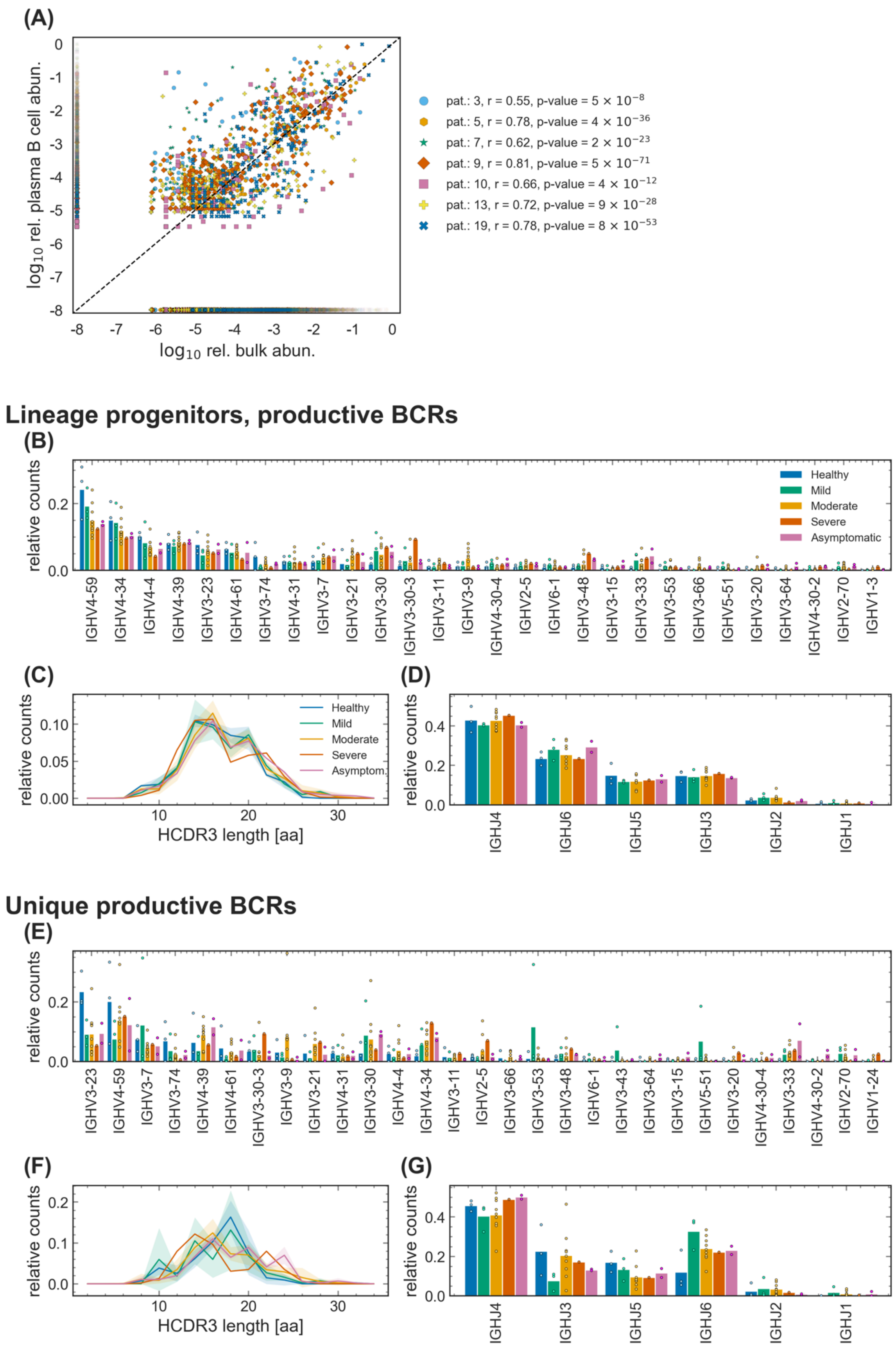

(A)
$\log_{10}$ rel. plasma B cell abun.
$\log_{10}$ rel. bulk abun.
pat.: 3, r = 0.55, p-value = $5 \times 10^{-8}$
pat.: 5, r = 0.78, p-value = $4 \times 10^{-36}$
pat.: 7, r = 0.62, p-value = $2 \times 10^{-23}$
pat.: 9, r = 0.81, p-value = $5 \times 10^{-71}$
pat.: 10, r = 0.66, p-value = $4 \times 10^{-12}$
pat.: 13, r = 0.72, p-value = $9 \times 10^{-28}$
pat.: 19, r = 0.78, p-value = $8 \times 10^{-53}$
Lineage progenitors, productive BCRs
(B)
relative counts
Healthy
Mild
Moderate
Severe
Asymptomatic
IGHV4-59 IGHV4-34 IGHV4-4 IGHV4-39 IGHV3-23 IGHV4-61 IGHV3-74 IGHV4-31 IGHV3-7 IGHV3-21 IGHV3-30 IGHV3-30-3 IGHV3-11 IGHV3-9 IGHV4-30-4 IGHV2-5 IGHV6-1 IGHV3-48 IGHV3-15 IGHV3-33 IGHV3-53 IGHV3-66 IGHV5-51 IGHV3-20 IGHV3-64 IGHV4-30-2 IGHV2-70 IGHV1-3
(C)
relative counts
HCDR3 length [aa]
Healthy
Mild
Moderate
Severe
Asymptom.
(D)
relative counts
IGHJ4 IGHJ6 IGHJ5 IGHJ3 IGHJ2 IGHJ1
Unique productive BCRs
(E)
relative counts
IGHV3-23 IGHV4-59 IGHV3-7 IGHV3-74 IGHV4-39 IGHV4-61 IGHV3-30-3 IGHV3-9 IGHV3-21 IGHV4-31 IGHV3-30 IGHV4-4 IGHV4-34 IGHV3-11 IGHV2-5 IGHV3-66 IGHV3-53 IGHV3-48 IGHV6-1 IGHV3-43 IGHV3-64 IGHV3-15 IGHV5-51 IGHV3-20 IGHV4-30-4 IGHV3-33 IGHV4-30-2 IGHV2-70 IGHV1-24
(F)
relative counts
HCDR3 length [aa]
(G)
relative counts
IGHJ4 IGHJ3 IGHJ5 IGHJ6 IGHJ2 IGHJ1

**Figure S3. Sequence features of immune receptors in the plasma B-cell repertoire across cohorts. (A)** Scatter plot shows $\log_{10}$ relative abundance of clonal lineages constructed from the plasma B-cell and bulk repertoire data from all time points and replicates in each patient (colors). To avoid primer-specific amplification biases, the relative abundance is estimated as the total read count of a clonal lineage relative to the total reads in the data associated with a specific primer amplification. Lineages with only bulk reads or only plasma reads are displayed as having $\log_{10}$ relative abundance = 1e-8. Pearson correlations (r) between abundances of lineages which were present in both the bulk and the plasma B-cell repertoires and the corresponding p-values are indicated in the legend for each patient. **(B-D)** Similar statistics are shown as in Fig. 3 (A,C,D), but for progenitors of clonal lineages with minimum size of three, in which at least one BCR is found in the plasma B-cell repertoire data; statistics of these lineages are reported in Tables S1, S2. Smaller read counts in the plasma B-cell data compared to the bulk do not allow for comparative analysis of receptor statistics across cohorts. **(D-F)** Similar statistics are shown as in Fig. S2 (A-C), but for unique receptors harvested from the plasma B-cell repertoires. Statistics of these receptors in each individual is described in Table S2. Smaller read counts in the plasma B-cell data compared to the bulk don’t allow for comparative analysis of receptor statistics across cohorts. Colors are consistent across panels.

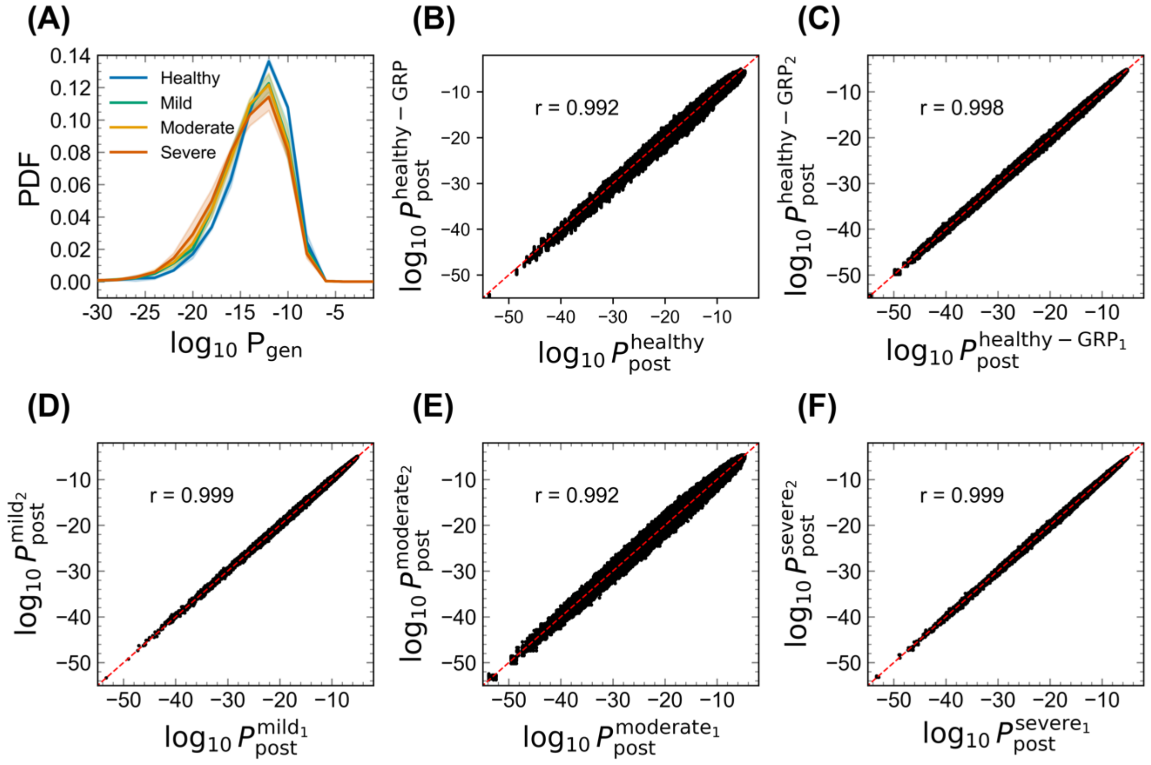


**Figure S4. Robustness of SONIA selection models.** **(A)** The distribution of the log-generation probability of a sequence $\sigma$ $\log_{10} \mathrm{P}_{\mathrm{gen}}(\sigma)$, evaluated using the inferred generation models by the IGoR software (Marcou et al., 2018), is shown as a normalized probability density function (PDF) for inferred naïve progenitors of productive clonal lineages in cohorts of healthy individuals and the mild, moderate, and severe cohorts of COVID-19 patients (colors). Full lines show distributions averaged over individuals in each cohort, and shadings indicate regions containing one standard deviation of variation among individuals within a cohort. **(B)** The scatterplot shows log $P_{\mathrm{post}}$ obtained by evaluating 500,000 generated sequences using the inferred selection (SONIA) models (Sethna et al., 2020) trained on the healthy cohort (x-axis) and 30 SONIA models trained on independent samples of the GRP dataset (Briney et al., 2019) down-sampled to the size of the healthy cohort in this study (7,161 receptors) (y-axis). The scatterplots show all unique pairwise comparisons between SONIA models trained on independent subsets with each cohort for **(C)** GRP (30 models), and COVID-19 patients with **(D)** mild (two models), **(E)** moderate (13 models), and **(F)** sever (three models) symptoms (Methods). The Pearson correlation between for pairwise model comparisons are shown in each panel.

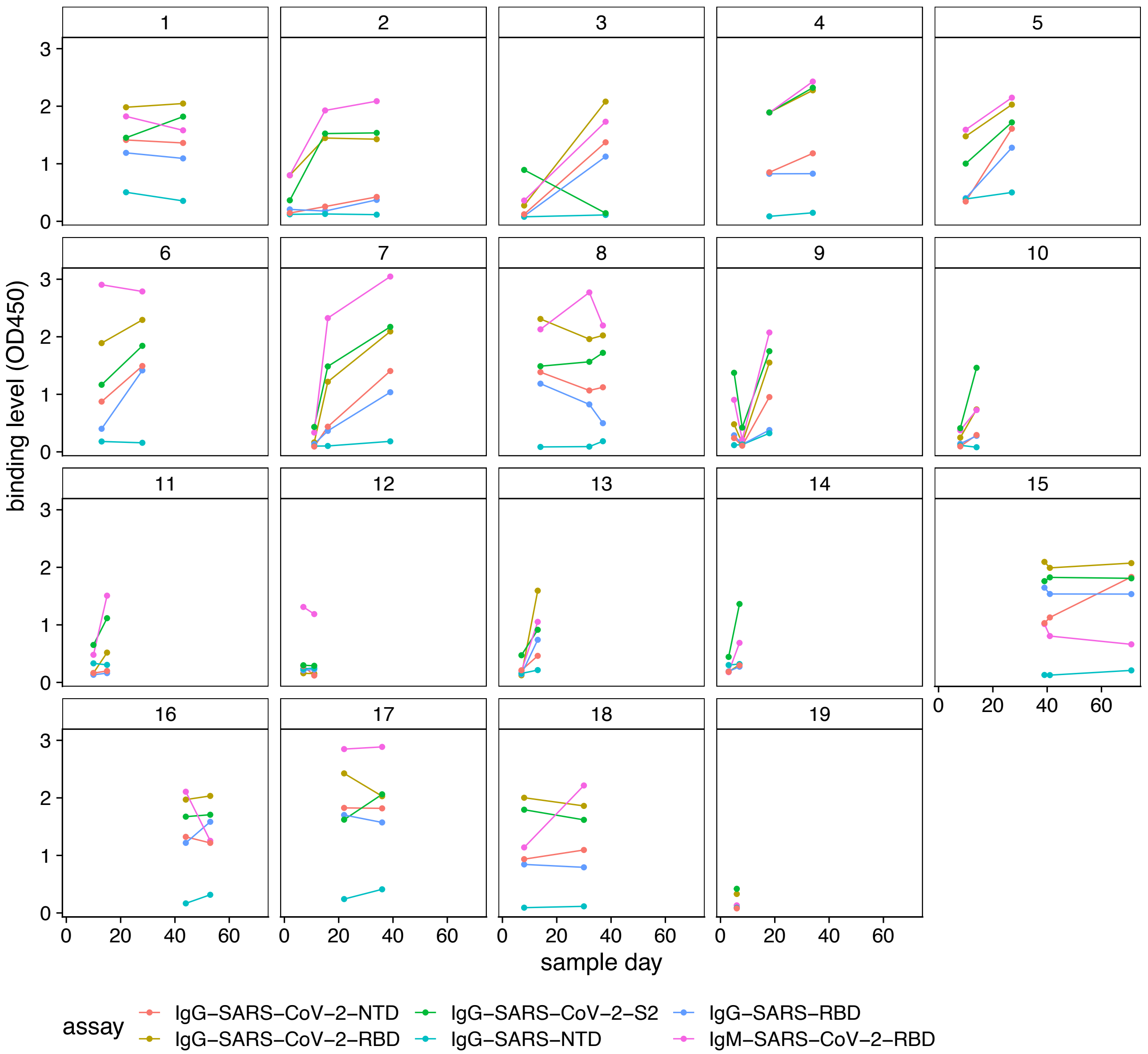


**Figure S5. ELISA binding assays for IgG and IgM repertoires against SARS-CoV-2 and SARS-CoV.** Plasma binding levels (measured by $OD_{450}$ in ELISA) against RBD, NTD, and S2 subdomain of SARS-CoV-2 and against RBD and NTD epitopes of SARS-CoV are shown. As seen in binding assays, many individuals developed a cross-reactive response to SARS-CoV-2 and SARS-CoV. Some individuals showed no increase in IgG binding to SARS-CoV-2 RBD due to already high levels at sampling time or natural variation. For the expansion analysis (Fig. 4), we analyzed only individuals whose IgG repertoires showed an increase in binding to SARS-CoV-2 (RBD): 2, 3, 4, 5, 6, 7, 9, 10, 11, 13, and 14.

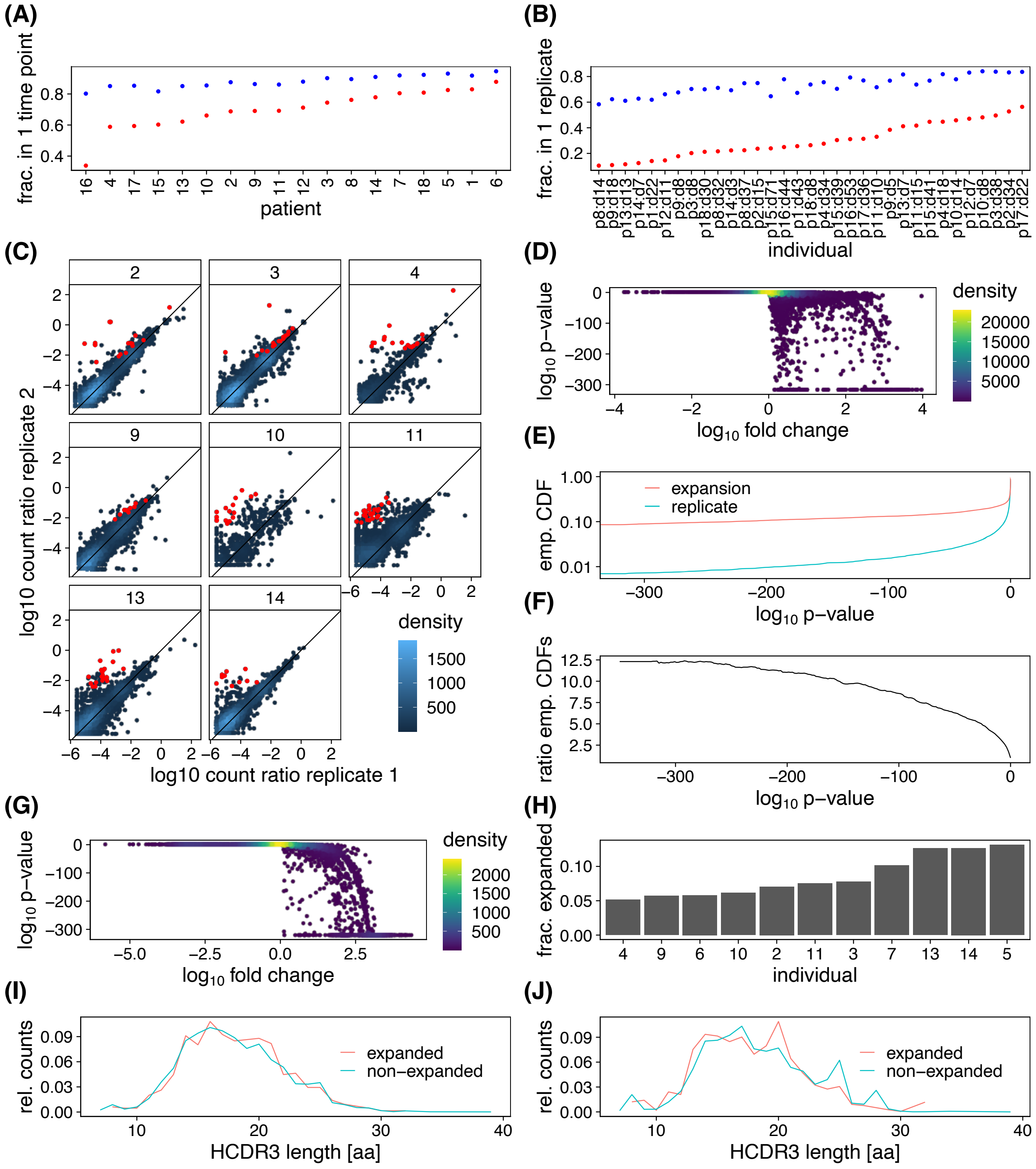


**Figure S6. Expansion supplement.** BCR repertoires are highly under-sampled, and relatively few BCR lineages appear in multiple time points and replicates. **(A)** Fraction of lineages present in only one time point before (blue) and after (red) filtering out small lineages (i.e., those with less than three unique sequences per time point) are shown. **(B)** Fraction of lineages present in only one replicate before (blue) and after (red) filtering out small lineages (i.e., those with less than three unique sequences per time point) are shown. **(C)** The log-ratio of abundance of receptors for all clonal lineages present in two replicates are shown. Each panel shows the test result for a given patient, as indicated in the label. The count density indicates the number of lineages at each point. Lineages that show a significant expansion over time are indicated in red. Since this is replicate data and represents a null model, red points indicate false positives. **(D)** $\log_{10}$ p-values of the expansion test versus $\log_{10}$ fold change (or odds ratio) for replicate data are shown. Color indicates density of points, and p-values of zero are displayed at the minimum nonzero value. See Methods for normalization, data processing, and hypothesis test. **(E)** Empirical cumulative density functions (CDF) of expansion data from multiple time points (red) and replicate data (blue) show that many more tests in expansion data result in low p-values compared to replicate data. **(F)** Ratio of empirical cumulative density functions (CDF) indicates that at a significance threshold of $10^{-300}$ there are roughly 12.3 times more positives than false positives. **(G)** $\log_{10}$ p-values of the expansion test versus $\log_{10}$ fold change (or odds ratio) for data corresponding to Fig. 4B is shown. Color indicates density of points, and p-values of zero are displayed at the minimum nonzero value. See Methods for normalization, data processing, and hypothesis test. **(H)** Fraction of lineages expanded for different individuals is shown. HCDR3 length distributions of expanded and non-expanded lineages, **(I)** with each lineage having equal weight, and **(J)** with each lineage weighted by the number of unique sequences per time point (excluding singletons) are shown.

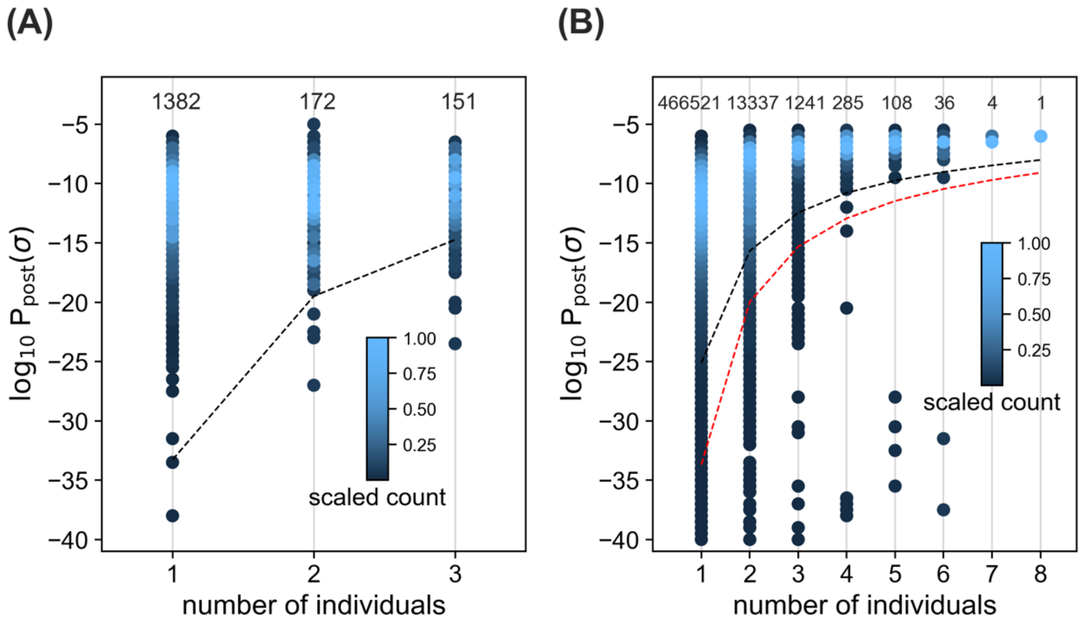


**Figure S7. Sharing of BCRs among healthy individuals. (A)** The density plot shows the distribution of $\log_{10} P_{\text{post}}$ for progenitors of clonal lineages shared in a given number of healthy individuals, indicated on the horizontal axis; histogram bin size is 0.5. The clonal lineages are constructed from the bulk data (Tables S1). The counts in each bin are scaled such that the maximum is equal to one for each column. The numbers above each column indicate the total number of sequences in the respective column. Sharing of rare lineages with $\log_{10} P_{\text{post}}$ below the dashed line is statistically significant (see Methods). **(B)** Similar statistics as in **(A)** are shown but for healthy individuals in the Great Repertoire Project (Briney et al., 2019). Sharing of rare lineages with $\log_{10} P_{\text{post}}$ below the black dashed line is statistically significant (see Methods). For comparison, the dashed line in **(A)** is shown as a red dashed line in **(B)** and extended to eight individuals.

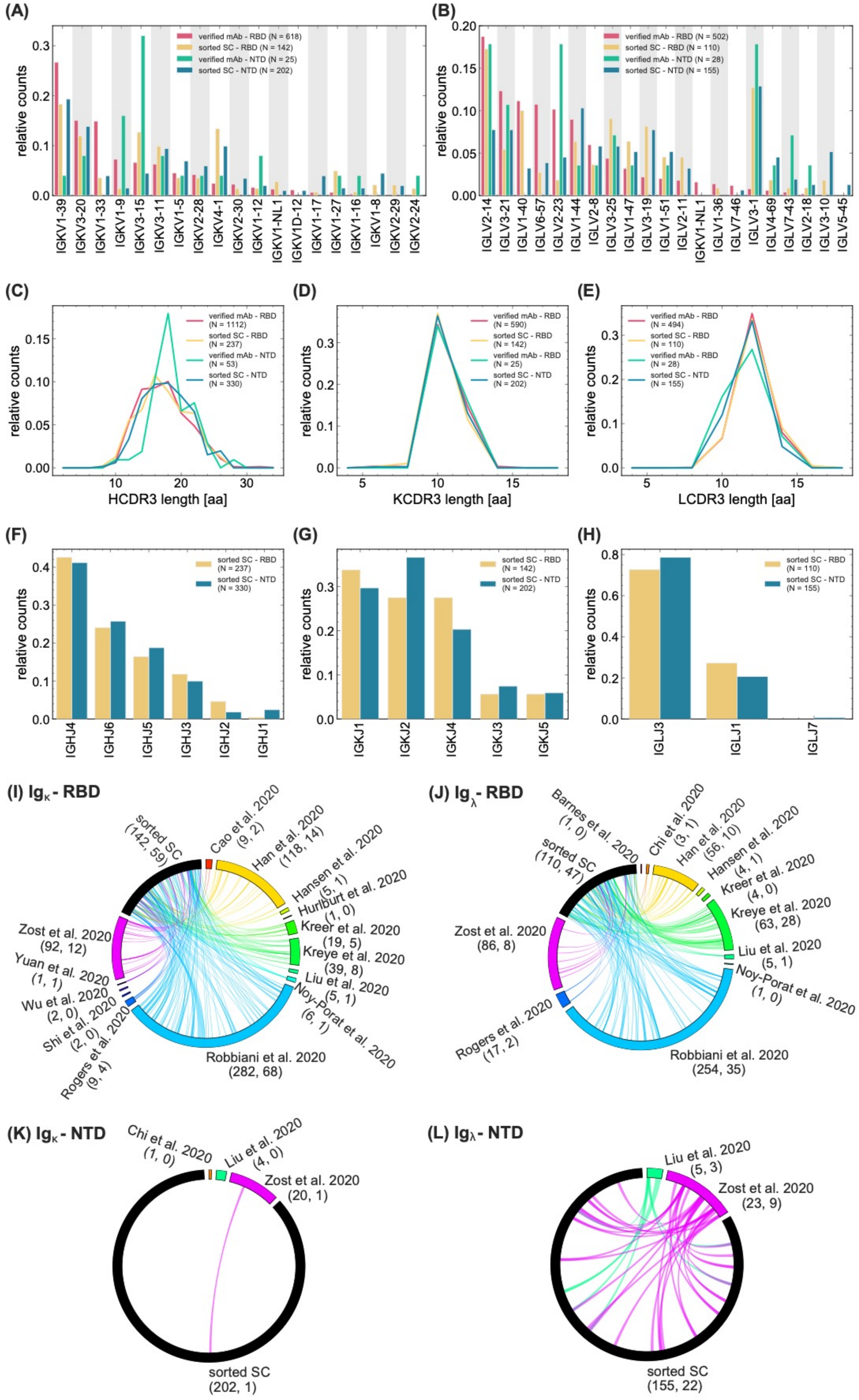

(A)
verified mAb - RBD (N = 618)
sorted SC - RBD (N = 142)
verified mAb - NTD (N = 25)
sorted SC - NTD (N = 202)
relative counts
IGKV1-39
IGKV3-20
IGKV1-33
IGKV1-9
IGKV3-15
IGKV3-11
IGKV1-5
IGKV2-28
IGKV4-1
IGKV2-30
IGKV1-12
IGKV1-NL1
IGKV1D-12
IGKV1-17
IGKV1-27
IGKV1-16
IGKV1-8
IGKV2-29
IGKV2-24
(B)
verified mAb - RBD (N = 502)
sorted SC - RBD (N = 110)
verified mAb - NTD (N = 28)
sorted SC - NTD (N = 155)
relative counts
IGLV2-14
IGLV3-21
IGLV1-40
IGLV6-57
IGLV2-23
IGLV1-44
IGLV2-8
IGLV3-25
IGLV1-47
IGLV3-19
IGLV1-51
IGLV2-11
IGKV1-NL1
IGLV1-36
IGLV7-46
IGLV3-1
IGLV4-69
IGLV7-43
IGLV2-18
IGLV3-10
IGLV5-45
(C)
verified mAb - RBD (N = 1112)
sorted SC - RBD (N = 237)
verified mAb - NTD (N = 53)
sorted SC - NTD (N = 330)
relative counts
HCDR3 length [aa]
(D)
verified mAb - RBD (N = 590)
sorted SC - RBD (N = 142)
verified mAb - RBD (N = 25)
sorted SC - NTD (N = 202)
relative counts
KCDR3 length [aa]
(E)
verified mAb - RBD (N = 494)
sorted SC - RBD (N = 110)
verified mAb - RBD (N = 28)
sorted SC - NTD (N = 155)
relative counts
LCDR3 length [aa]
(F)
sorted SC - RBD (N = 237)
sorted SC - NTD (N = 330)
relative counts
IGHJ4
IGHJ6
IGHJ5
IGHJ3
IGHJ2
IGHJ1
(G)
sorted SC - RBD (N = 142)
sorted SC - NTD (N = 202)
relative counts
IGKJ1
IGKJ2
IGKJ4
IGKJ3
IGKJ5
(H)
sorted SC - RBD (N = 110)
sorted SC - NTD (N = 155)
relative counts
IGLJ3
IGLJ1
IGLJ7
(I) Igκ - RBD
sorted SC (142, 59)
Cao et al. 2020 (9, 2)
Han et al. 2020 (118, 14)
Hansen et al. 2020 (5, 1)
Hurlburt et al. 2020 (1, 0)
Kreer et al. 2020 (19, 5)
Kreye et al. 2020 (39, 8)
Liu et al. 2020 (5, 1)
Noy-Porat et al. 2020 (6, 1)
Robbiani et al. 2020 (282, 68)
Rogers et al. 2020 (9, 4)
Shi et al. 2020 (2, 0)
Wu et al. 2020 (2, 0)
Yuan et al. 2020 (1, 1)
Zost et al. 2020 (92, 12)
(J) Igλ - RBD
Barnes et al. 2020 (1, 0)
sorted SC (110, 47)
Chi et al. 2020 (3, 1)
Han et al. 2020 (56, 10)
Hansen et al. 2020 (4, 1)
Kreer et al. 2020 (4, 0)
Kreye et al. 2020 (63, 28)
Liu et al. 2020 (5, 1)
Noy-Porat et al. 2020 (1, 0)
Robbiani et al. 2020 (254, 35)
Rogers et al. 2020 (17, 2)
Zost et al. 2020 (86, 8)
(K) Igκ - NTD
Chi et al. 2020 (1, 0)
Liu et al. 2020 (4, 0)
Zost et al. 2020 (20, 1)
sorted SC (202, 1)
(L) Igλ - NTD
Liu et al. 2020 (5, 3)
Zost et al. 2020 (23, 9)
sorted SC (155, 22)

**Figure S8. Sequence features of heavy and light chain receptors in sorted single cells and monoclonal antibodies.** The bar graphs show the relative counts for **(A)** the $\kappa -$ chain IGKV-gene usage and **(B)** the $\lambda -$ chain IGLV-gene usage for the verified mAbs reactive to RBD (pink) and NTD (green) epitopes of SARS-CoV-2 (Table S7) and the light chain receptors obtained from the RBD- (yellow) and NTD- (blue) sorted single cell data (Methods). Distributions of the lengths of **(C)** of HCDR (heavy chain), **(D)** KCDR3 ($\kappa -$ chain), and **(E)** LCDR3 ($\lambda -$ chain ) amino acid sequences are shown. **(F)** IGHJ-gene usage, **(G)** IGKJ-gene usage, and **(H)** IGLJ-gene usage of the sorted single cells is shown in relative counts in bar graphs. Colors are consistent between panels and the number of samples used to evaluate the statistics in each panel is indicated in the legend. **(I-L)** Circos plots show matches between the light chain CDR3 sequences of progenitors in the sorted single cell dataset (black) and light chain CDR3 sequences in the verified mAbs (colors) for RBD-reactive **(I)** $\mathrm{IG}_{\kappa}$, and **(J)** $\mathrm{IG}_{\lambda}$ sequences, and for NTD-reactive **(K)** $\mathrm{IG}_{\kappa}$, and **(L)** $\mathrm{IG}_{\lambda}$ sequences. Different colors indicate different studies from which mAbs were pooled. The reference to each study, the total number of mAbs in the study, and the number of mAbs with matching light chain CDR3 to the single cell data are reported in each panel.

TableS1.xlsx

**Table S1. Statistics of BCR repertoire sequence data from healthy individuals, and COVID-19 patients.** The information about individuals in each cohort is shown. Detailed statistics of the processed data for productive BCRs are shown for read abundance, number of singletons, and number of unique sequences for all replicates and sampled time-points in each individual. For each individual, the number of lineages with more than two and ten unique sequences across all time points are shown in separate columns. Read statistics for unproductive receptors pooled from all individuals are shown separately.

| 5'-end primer | Sequence (5'-3') |
|---|---|
| IGHV1 | CCTCAGTGAAGGTCTCCTGCAAGG |
| IGHV2 | TCCTGCGCTGGTGAAACCCACACA |
| IGHV3 | GGTCCCTGAGACTCTCCTGTGCA |
| IGHV4 | TCGGAGACCCTGTCCCTCACCTGC |
| IGHV5 | CAGTCTGGAGCAGAGGTGAAA |
| IGHV6 | CCTGTGCCATCTCCGGGGACAGTG |
| **3'-end primer** | **Sequence (5'-3')** |
| CHG-R | GCGCCTGAGTTCCACGACAC |

**Table S2. List of primers used for PCR amplification of B-cell repertoires samples.**

| Patient (Sex, Age) | Unique Sequences | Lineages (size > 2) | Lineages (size > 10) |
|---|---|---|---|
| 316188 (F, 30) | 204218 | 13138 | 3202 |
| 326650 (F, 18) | 572194 | 28785 | 6139 |
| 326651 (M, 18) | 6174253 | 180993 | 54272 |
| 326713 (F, 25) | 3087960 | 104242 | 29682 |
| 326780 (M, 29) | 297727 | 15195 | 3850 |
| 326780 (M, 29) | 526166 | 26392 | 5039 |
| 326797 (F, 21) | 1538645 | 56487 | 11467 |
| 326907 (F, 29) | 73418 | 4555 | 1124 |
| 327059 (M, 26) | 1932407 | 64786 | 20140 |
| D103 (M, 25) | 169950 | 8423 | 2745 |
| Pooled unproductive | 166704 | 11534 | 1049 |

**Table S3. Statistics of IgG BCR repertoire sequence data from individuals in the Great Repertoire Project.** Because of the massive amount of data provided by the Great Repertoire Project (Briney et al., 2019), only the first three biological replicates were used for each individual to be comparable to the data sampled in this study. Detailed statistics of the processed data for productive BCRs are shown for the number of unique sequences pooled from all replicates for each individual. For each individual, the number of lineages with more than two and ten unique sequences are shown in separate columns. Read statistics for unproductive receptors pooled from all individuals and replicates are shown separately.

**TableS4.xlsx**

**Table S4. Statistics of plasma B-cell repertoire sequence data from COVID-19 patients.** The information about individuals in each cohort is shown. Detailed statistics of the processed data for productive BCRs are shown for read abundance, number of singletons, and number of unique sequences for all replicates and sampled time points in each individual. For each individual, the amounts of lineages with more than two and ten unique sequences across all time points are shown in separate columns which are split further by whether the lineages also contained bulk reads.

**TableS5.xlsx**

**Table S5. Rare expanding BCRs shared among individuals.** The list of 38 rare progenitors of clonal lineages (i.e., with $P_{\text{post}}$ below the dashed line in Fig. 6) that exhibit lineage expansion in at least one individual is shown. These receptors are indicated by green diamonds in Fig. 6. The presence of these lineages in the plasma B-cell repertoire is indicated in the last column (orange triangles in Fig. 6). The 38 rare expanding lineage progenitors shown here are shared among four to 12 COVID-19 patients.

**TableS6.xlsx**

**Table S6. Single-cell sorted antibodies matched with BCR repertoires of COVID-19 patients.** Receptors from RBD- and NTD-sorted single cell data whose HCDR3 sequences and IGHV-gene clustered with a BCR lineage constructed from the bulk+plasma B-cell repertoires of patients in this study are shown (Methods).

**TableS7.xlsx**

**Table S7. Verified antibodies detected in BCR repertoires of COVID-19 patients.** HCDR3 and IGHV-gene of verified monoclonal antibodies responsive to SARS-CoV-2 (RBD, NTD, and S1) or SARS-CoV-1 epitopes, whose HCDR3 sequences match with a receptor (with up to Hamming distance of one amino acid) in the bulk+plasma B-cell repertoires of patients in this study are shown. Each row indicates a monoclonal antibody family, whose members have similar HCDR3, up to one amino acid difference. Mutations in the repertoire-matched receptors with respect to the original HCDR3 are in red. Single amino acid mutation differences in HCDR3s of monoclonal antibody families are shown in cyan. Patient ID for each repertoire-matched receptor is indicated in the last column. The complete list of verified antibodies is given in Table S8.

**TableS8.xlsx**

**Table S8. Complete list of verified monoclonal antibodies.**
